# Microbial Community of Small Intestine in Acute Severe Pancreatitis Patients: a Pilot Study

**DOI:** 10.1101/2021.08.24.21262159

**Authors:** Vladimir V. Kiselev, Alexey V. Kurenkov, Sergey S. Petrikov, Petr A. Yartsev, Vera E. Odintsova, Stanislav I. Koshechkin, Alexander V. Tyakht

## Abstract

Purpose of the study: to describe the composition of the microbiota of the initial sections of the small intestine in patients with severe necrotizing acute pancreatitis.

**Objectives of the study:**

1. Determine the composition of the microbiota of the initial sections of the small intestine upon admission to the ICU;
2. Determine the differences in the composition of the microbiota of the initial sections of the small intestine, depending on the timing of the onset of the disease.

**Introduction:** Disturbance of intestinal homeostasis is a leading factor in the pathogenesis and progression of systemic inflammation in patients with severe acute pancreatitis. The development of systemic complications occurs due to both mesenteric hypoperfusion and dysregulation of intestinal motility, and the destruction of the intestinal barrier, with the translocation of bacterial bodies and their substrates. Which increases the risk of developing POI and increasing mortality. With the advent of methods for high-throughput sequencing of microbiome samples - for example, in the 16S rRNA format - the possibilities for studying the structure of microbial communities have significantly expanded. In this regard, there is more and more evidence of the relationship between the state of human health and microflora inhabiting various parts of the body.

**Materials and methods:** The study included 7 patients with a diagnosis of severe necrotizing acute pancreatitis (6 men, 1 woman), the mean age was 54.1 ± 14.4 years. The patients were divided into two groups. Group 1 (n = 4) included patients admitted 2-4 days after the onset of a pain attack. Group 2 (n = 3) - patients admitted no later than 24 hours from the onset of the disease. The bacterial composition of jejunal wash samples was studied using 16S RNA sequencing. The severity of the condition was assessed using the integral scales APACHE II, SOFA, SAPS II. In patients of the main group, APACHE II was 22 ± 2.83 points (18; 24), SOFA - 6.8 ± 0.5 points (6; 7), SAPSII - 32.9 ± 6.4 points (24.7; 40), in patients of the comparison group, APACHE II is 18.0 ± 3.7 points (12; 22), SOFA - 4.0 ± 2.6 points (2; 7), SAPSII - 24.4 ± 5.0 points (20.9; 30.1).

Material was collected at the time of installation of a sterile multifunctional intestinal catheter for Treitz’s ligament, no later than 12 hours from the moment of admission to the ICU. At the time of sampling, patients were not receiving antibiotic therapy.

**Results:** A more severe course was associated with a reduced representation in the microbiome of the species *Nesseria* mucosa and *Parvimonas* micra inhabiting the mucosal layer, as well as *Megasphaera micronuciformis*. The share of *Streptococcus* genera (*S. rubneri / parasanguinis / australis* species) and *Actinomyces* and a number of genera from the *Enterobacteriaceae* family in such patients, on the contrary, was higher.

**Interest disclosure:** Sample preparation, sequencing and analysis of these microbiome samples was carried out by Knomics LLC during a commercial project for VneshPromFarm LLC, the manufacturer of saline electrolyte solution (SES).

## Introduction

Disruption of intestinal homeostasis is a leading factor in the pathogenesis and progression of systemic inflammatory response in patients with acute severe pancreatitis. In turn, systemic complications develop both as a result of mesenteric hypoperfusion and dysregulation of intestinal motility, and the destruction of the intestinal barrier, with the translocation of bacterial bodies and their substrates. It should be noted that all of the above processes increase the risk of developing multiple organ failure, leading to an increase in the number of deaths [1]. With the advent of methods for high-throughput sequencing of microbiome samples - for example, in the 16S rRNA format - the possibilities for studying the structure of microbial communities have significantly expanded. In this regard, there is more and more evidence of the relationship between the state of human health and the microflora inhabiting various parts of his body [2].

The commensal microflora in the small intestine of a healthy person is approximately 103-104 different types of bacteria. They are indispensable for the digestion of food substrates, participate in the generation of secondary bile acids, which promote the absorption of dietary lipids and fat-soluble vitamins [3], have a proliferative effect on intestinal epithelial cells, promote the differentiation of enterocytes, prevent colonization by pathogenic microorganisms and form a local and systemic immune response [4]. As a result of the fermentation of complex carbohydrates, short-chain fatty acids are formed, which serve as an energy source for the macroorganism and confer beneficial effects on immune cells [5], proliferation, differentiation and function of intestinal epithelial cells [6], and participate in the regulation of metabolism [7]. Thus, the intestinal microbiota and its metabolic products are vital for intestinal homeostasis.

Numerous clinical and experimental studies demonstrate that pancreatic ischemia plays an important role in the development of acute pancreatitis and disease progression with further development of pancreatic necrosis [8-10]. Microcirculation disorders are caused by a decrease in capillary blood flow in the tissues of the pancreas against the background of inflammation. Hypotension associated with systemic inflammatory response syndrome usually leads to centralization of blood circulation due to shunting of blood from peripheral vessels into the main bloodstream. Microcirculatory disorders involve the small intestine, resulting in neuroendocrine dysregulation and epithelial dysfunction of enterocytes, which are an important source of proinflammatory mediators such as IL-17, lipid mediators produced by phospholipase A2, and antimicrobial peptides [11,12]. At the same time, an increase in the proportion of pathogenic microorganisms in the microbiota of the small intestine leads to a syndrome of bacterial overgrowth and translocation of microbial substrates. Bacterial translocation and bacteremia lead to further escalation of systemic inflammation, and in some cases, to the development of sepsis, septic shock and circulatory failure, with multiple organ dysfunction syndrome [11].

Diagnosis and treatment of intestinal failure syndrome (AIC) is difficult task, since intestinal dysfunction and gastrointestinal tract insufficiency develop in the most severe patients with a long stay in the intensive care unit (ICU) [13]. Instrumental methods for detecting manifestations of SCI remain complex, and laboratory markers do not always reflect the real picture of critical events in the small intestine [1,14–16]. With the advent of methods for high-throughput sequencing of microbiome samples - for example, in the 16S rRNA format - the possibilities for studying the structure of microbial communities have significantly expanded. In this regard, there is more and more evidence of the relationship between the state of human health and the microflora inhabiting various parts of his body. DNA analysis of the microbiome provides new opportunities for early correction of intestinal microbiota disorders and prevention of the development of AIC, including in critically ill patients.

At the moment, there is data on the colon microbiome in patients with varying degrees of severity of pancreatitis [17]. However, a more accurate picture of changes can be expected in the microbiota of the small intestine. This paper investigates the microbial composition from the jejunum. Samples were obtained from 7 patients with acute pancreatitis with varying degrees of severity. Including a comparison of two samples taken from one patient with a difference of 24 hours against the background of an enteral infusion with saline electrolyte solution (SES).

## Materials and Methods

### Ethic statements

Ethical approval for the study was obtained from the local ethic committee of Sklifosovsky Research Institute of Emergency Care of the Moscow Healthcare Department. We declare that all methods were performed in accordance with the relevant guidelines and regulations. All patients gave a written informed consent for the endoscopy.

### Patient enrollment and washing sample collection

The study included 7 patients (6 men, 1 woman, age 54.1 +/- 14.4 years (43; 76)) with severe necrotizing acute pancreatitis, confirmed by CT and ultrasound studies. The etiological cause of the development of this disease was the alcohol factor. All patients had abdominal pain, nausea, vomiting, increased levels of pancreatic enzymes in serum.

The patients were divided into two groups: the first group (n = 4, main) included patients who were admitted to 2 4 days from the onset of the disease (from the onset of a painful attack). The second group (n = 3, comparisons) included patients who were admitted no later than 24 hours from the onset of the disease. The severity of the condition was assessed by APACHE II, SOFA, SAPS II. The patients of the main group admitted on days 2-4 from the onset of the disease had higher indices on the integral scales of severity: APACHE II - 22 ± 2.83 points (18; 24), SOFA - 6.8 ± 0.5 points (6; 7), SAPSII - 32.9 ± 6.4 points (24.7; 40), compared with patients in the comparison group, APACHE II 18.0 ± 3.7 points (12; 22), SOFA - 4, 0 ± 2.6 points (2; 7), SAPSII - 24.4 ± 5.0 points (20.9; 30.1).

Samples of small intestine washing were collected from each subject. Sample collection was performed during intubation by multifunctional intestinal catheter (utility model patent application “Multifunctional intestinal catheter”, RU 199398 U1; https://patents.google.com/patent/RU199398U1/ru) beyond the ligament of Treitz not later than 12 hours past the admission to the ICU. From one of the main group of patients, two samples were collected with a 24 hours interval. The patient’s catheter got dislocated and the second sample was collected during the re-intubation. Intubation and sample collection were performed with EXERA II SIF-Q180 small intestinal videoscope (Olympus) (utility model patent application “Method of biologic fluid collection from the small intestine through an endoscopic channel”, RU 2738007 C1; https://patents.google.com/patent/RU2738007C1/ru?oq=RU2738007C1). The catheter had an original construction. In brief, it was a tube with an obturator on the distal end and a syringe on the proximal end. When the distal end of the catheter reached the destination, the obturator was pushed out by physiological salt solution flow. The construction excluded the chance of probe contamination by the microorganisms from other biological media and provided the ability to collect a sufficient amount of biomaterial for further 16S rRNA gene sequencing.

The SES for enteral infusion (patent RU 2 699 22 C1 dated 2019.09.14; https://patents.google.com/patent/RU2699222C1/ru) that includes pectin, inulin and glutamine, was infused by the intestinal tube into each patient in order to stimulate intestinal motility. The SES infusion was performed at the speed of 6-10 ml/min, the volume was 1500 (±412.3) ml upon condition the abdominal pressure (AP) did not exceed 16-20 mm Hg. The AP measurement was performed according to the methodology recommended by World Society of the Abdominal Compartment Syndrome (WSACS).

### Microbiome analysis

DNA was extracted from the samples using DNeasy PowerLyzer Microbial kit (Qiagen). The sequencing library preparation was performed for a variable region V4 of 16S rRNA as described previously [18]. Sequencing was performed on Illumina MiSeq platform. The sequencing data were analyzed in the Knomics-Biota system [19]. Taxonomic profiling was performed using amplicon-sequence variants with DADA2 software [20]. Additional taxonomic classification of selected microbes was performed using the BLAST algorithm against NCBI nr database (https://blast.ncbi.nlm.nih.gov/Blast.cgi). If the whole sequence was completely identical to the reference sequences of multiple taxa, all of them were listed in its designation (for example, *Streptococcus rubneri/parasanguinis/australis*). Alpha diversity was obtained after 5-fold rarefaction to 3000 reads and averaging of the results for each iteration. Pairwise dissimilarity between samples was estimated with Aitchison distance calculated on unrarified reads.

In order to filter out the possible contamination of the samples during sample processing, the taxonomic composition was compared with the composition of three negative control samples (the blank clear water samples prepared and sequenced in parallel). Overall, the contamination was negligible. For further analysis, the taxa that were abundant in negative control and rare in the samples of interest were excluded. Several taxa detected in the negative control samples were preserved as they are known as inhabitants of the intestinal tract. The results of analysis are available in the Knomics-Biota at https://biota.knomics.ru/acute-severe-pancreatitis-pilot.

Additional analysis was conducted using R programming language [21] to determine the inter-group differences. The clr-transformation was applied to the relative abundance values of taxa[22]. According to this approach, the relation between taxa proportion in metagenome to certain averaged taxa proportion in logarithmic scale are analyzed instead of the relative abundances themselves. This approach provides a more correct estimate of differences between samples [23]). Only the taxa that were present in all samples of at least one group (except for SUU02, the second sample of a patient collected 24 hours after SES infusion) were included in the analysis. Replacements of zero values were imputed using the cmultRepl function from zCompositions library in R [24]. In order to identify the taxa that were the most discriminating between the groups, we selected the taxa for which the clr-transformed abundance was higher in all samples of one of the groups compared to all samples of the other group (except for the sample SUU02). Fisher’s exact test with Benjamini-Hochberg adjustment for multiple testing was used to compare taxa abundances in two samples of one patient. The analysis was performed at the levels of genus and unique sequences. Taxonomy of sequences was further refined with the BLAST algorithm against NCBI nr database.

### Availability of data and materials

The reads generated in the study were deposited in the Sequence Read Archive (SRA), project ID: PRJNA705601.

## Results

### Microbial composition of intestinal washing samples

Total taxonomic composition of the washing samples is shown on Figure 1. The Streptococcus was the most abundant genus overall. It was represented by a wide range of species, including *S. oralis/cristatus/mitis/infantis/gordonii, rubneri/parasanguinis/australis, anginosus, mutans, thermophilus/salivarius/vestibularis* and closely related unclassified species. These taxa belong to the normal microbiome of upper regions of a human digestive tract including small intestine [25]. Most of them belong to the *S. mitis* group. Recent studies provide evidence of association between decrease of antibodies to *S. mitis* in blood plasma and pancreatic cancer [26].

**FIGURE 1.**
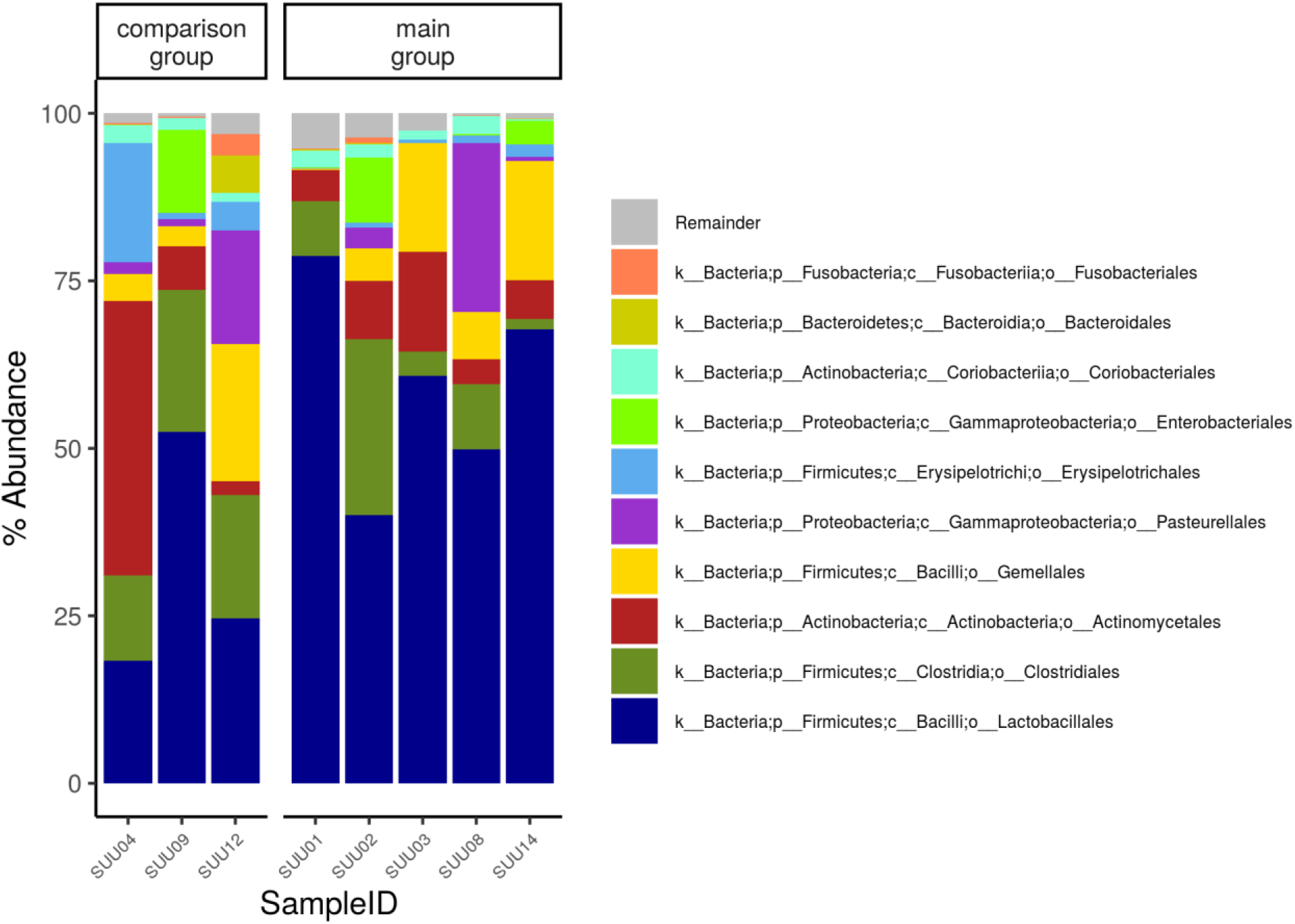
Taxonomic composition of samples at the order level. Sample SUU02 is the second sample of a main group patient. The first sample of this patient is SUU01

The unclassified bacteria from the *Gemellaceae* family were observed in all samples. Their taxonomic classification was refined to be *G. taiwanensis/parahaemolysans/sanguinis/morbillorum/haemolysans* by BLAST algorithm against NCBI database. These species are commensal inhabitants of a small intestine [25].

The *Peptostreptococcus* and *Solobacterium* were also present in each sample. According to the NCBI database, they were represented by *P. stomatis* and *S. moorei* species, respectively. Increased presence of these species in stool is associated with colorectal cancer [27].

The *Haemophilus* genus was represented by the *H. haemolyticus* and *parainfluenzae* species. *H. haemolyticus* is associated with chronic obstructive pulmonary disease [28] in several cases it has been found in the blood of patients with bacteremia [29]. *H. parainfluenzae* is mostly known as an infectious agent of lungs and upper airways. It may cause bacteremia and infective endocarditis. It has been found in microbiota analysis of patients with several diseases, including irritable bowel syndrome [30] and stomach cancer [31]. *Haemophilus* was observed in negative control samples at a small proportion.

The *Corynebacterium* was also among the highly abundant genera: its proportion in one of the comparison group samples was 35.9%. According to the NCBI database, the taxon was refined to be *C. argentoratense* or closely related species. This microbe was initially identified in the upper airways [32] and later in other parts of the human body, inter alia associated with infections [33].

A sample of one of the main group patients contained a high proportion of *Enterococcus* (17.3%). Some species of this genus are opportunistic pathogens, and high abundance of such species is not common for healthy human gut microbiota.

Microbiota of several patients contained unclassified species of *Enterobacteriaceae* family; although the sequenced gene fragment does not allow to distinguish the genera reliably, the family is known to include some opportunistic pathogens among *the Escherichia/Shigella* and *Klebsiella* genera.

Patients samples also contained such representatives of commensal microbiome as *Rothia* (*including R. mucilaginosa and dentocariosa*), *Granulicatella, Parvimonas, Veillonella* and *Actinomyces* (refined to be *A. odontolyticus*).

The Shannon index describing evenness of taxa proportions varied between 2.73 and 4.4. The Chao index characterizing diversity of microbiota with emphasis on proportion of rare taxa was between 72.32 and 132.01 (Figure 2). Aitchison distance between samples varied from 5.46 to 21.56.

**FIGURE 2.**
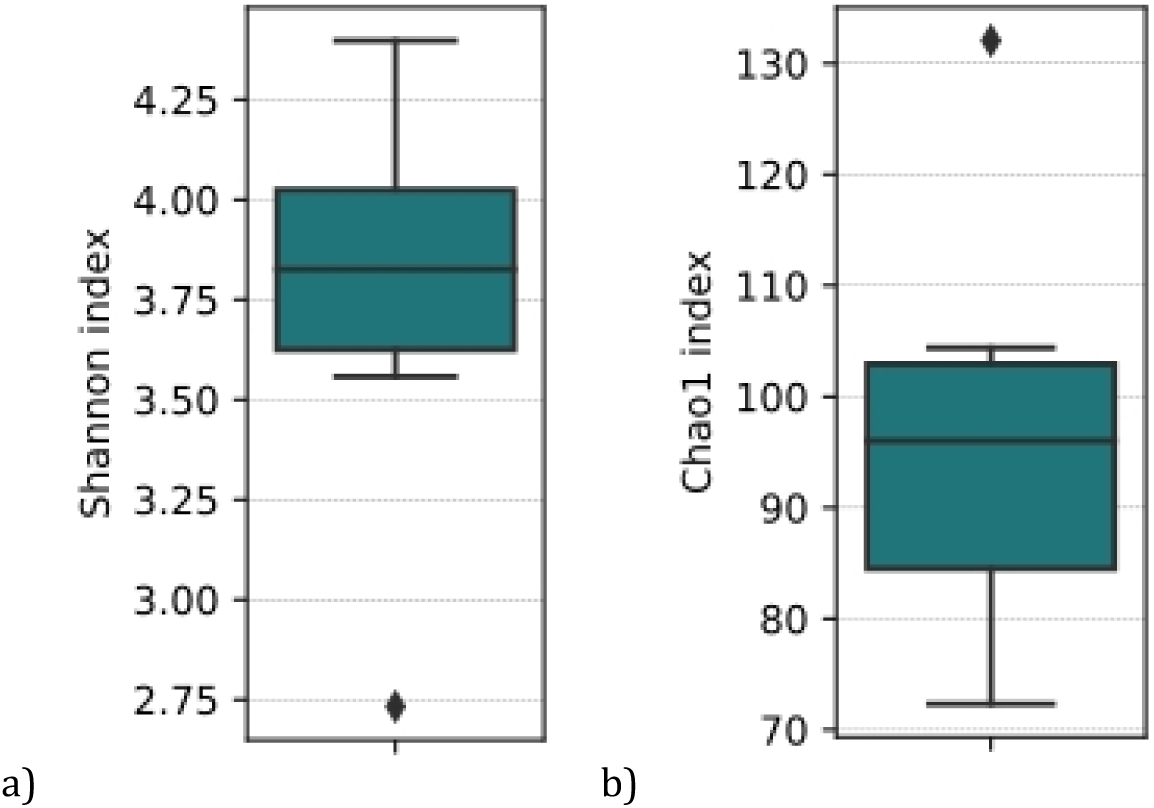
Alpha diversity of small intestinal washing samples for patients with acute pancreatitis: A, Shannon index; B, Chao1 index

### Microbial composition of intestinal washing samples is linked to clinical status

Patients of the main group (those were admitted to hospital on the day 2-4 after the disease onset) had higher points according to integrative severity scales: APACHE II - 22±2.83 points (18; 24), SOFA – 6.8±0.5 points (6; 7), SAPS II - 32.9±6.4 points (24.7; 40) - as compared to the comparison group: APACHE II 18.0±3.7 points (12; 22), SOFA – 4.0±2.6 points (2; 7), SAPS II - 24.4±5.0 points (20.9; 30.1).

The microbial composition of washing samples was also different for the main group as compared to the comparison group. The major difference among the highly-abundant taxa was observed for *Streptococcus rubneri/parasanguinis/australis*. It was found at larger proportions in microbiota of patients with higher scores according to the cumulative severity scales (Figure 3). These species correspond to the *S. mitis* group and are part of commensal intestinal and oral microbiome. Nevertheless, they may cause bacteremia, endocarditis and other diseases if they get into the bloodstream [34]. The opportunistic pathogens *Actinomyces* [35]and *Escherichia/Shigella* (or closely related *Brenneria/Pseudescherichia*) were also more abundant in microbiota of patients from the main group. Proportion of *Neisseria* mucosa, an inhabitant of intestinal mucus layer [36], and *Parvimonas* micra, which is specifically able to degrade mucin[37,38], was lower for the participants of the main group. Relative abundance of *Megasphaera* micronuciformis was also decreased in patients with higher disease severity [39,40].

**FIGURE 3.**
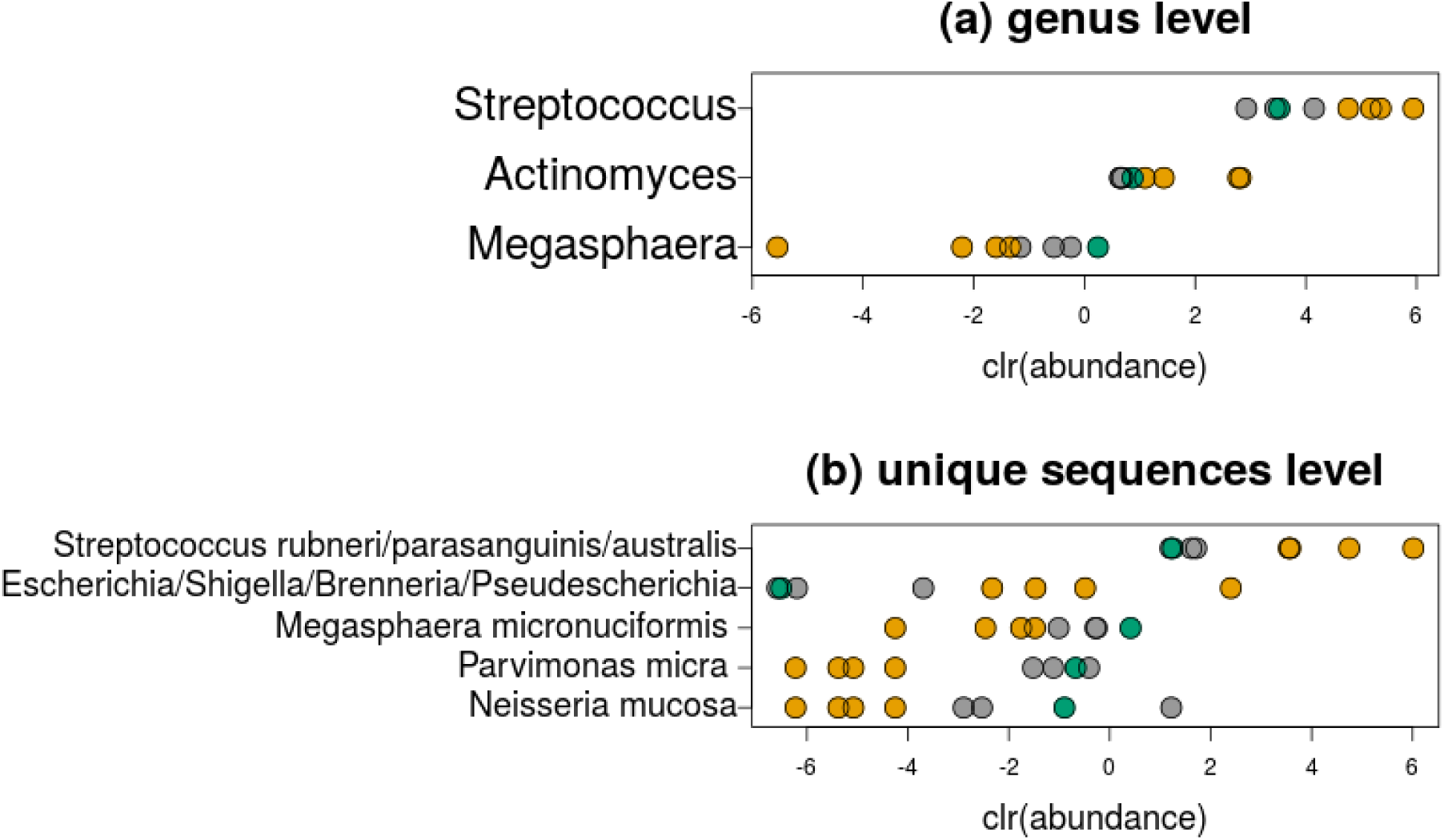
Clr-transformed relative abundance of several taxa at genus level and at the unique sequences level. Yellow color denotes samples of the comparison group, grey - samples of the main group. Green color is for sample SUU02 - the second sample of a patient from the main group.

### Comparison of two samples from a main group patient before and after SES infusion

For one of the main group patients, two samples were collected. The first one was obtained during the insertion of a sterile multifunctional intestinal catheter. The second one was collected 24 hours later, during the re-insertion due to the dislocation of the initially inserted catheter to the stomach.

In this sample, the abundance of the taxa listed in the previous section was closer to the abundance in the comparison group samples than to the values for the main group samples (Figure 3). *Parvimonas* micra, *Neisseria* mucosa and *Megasphaera* micronuciformis were not detected in the first sample of the patient and were found in a notable amount in the second one (*N. mucosa* - 0.34%, *P. micra* - 0.43%, *M. micronuciformis* - 1.3%). The *Streptococcus rubneri/parasanguinis/australis* relative abundance has reduced (from 24.58% to 2.93%), as well as the proportion of the whole *Streptococcus* genus (from 46.09% to 34.73%), *Actinomyces* genus (from 3.69% to 2.45%) and *Escherichia/Shigella/Brenneria/Pseudescherichia* (from 0.36% to 0%). In order to exclude the possibility that the detected difference was due to the variability in sequencing depth of the samples, Fisher’s exact test was performed for each of the bacteria. Results of the test provide support that the differences between two samples are not arbitrary (p<0.05).

As for beta-diversity (Aitchison distance), the second sample of the patient was also closer to the comparison group rather than to the main one (Figure 4). It was more similar to the samples of patients with lower severity than to the first sample of the subject.

**FIGURE 4.**
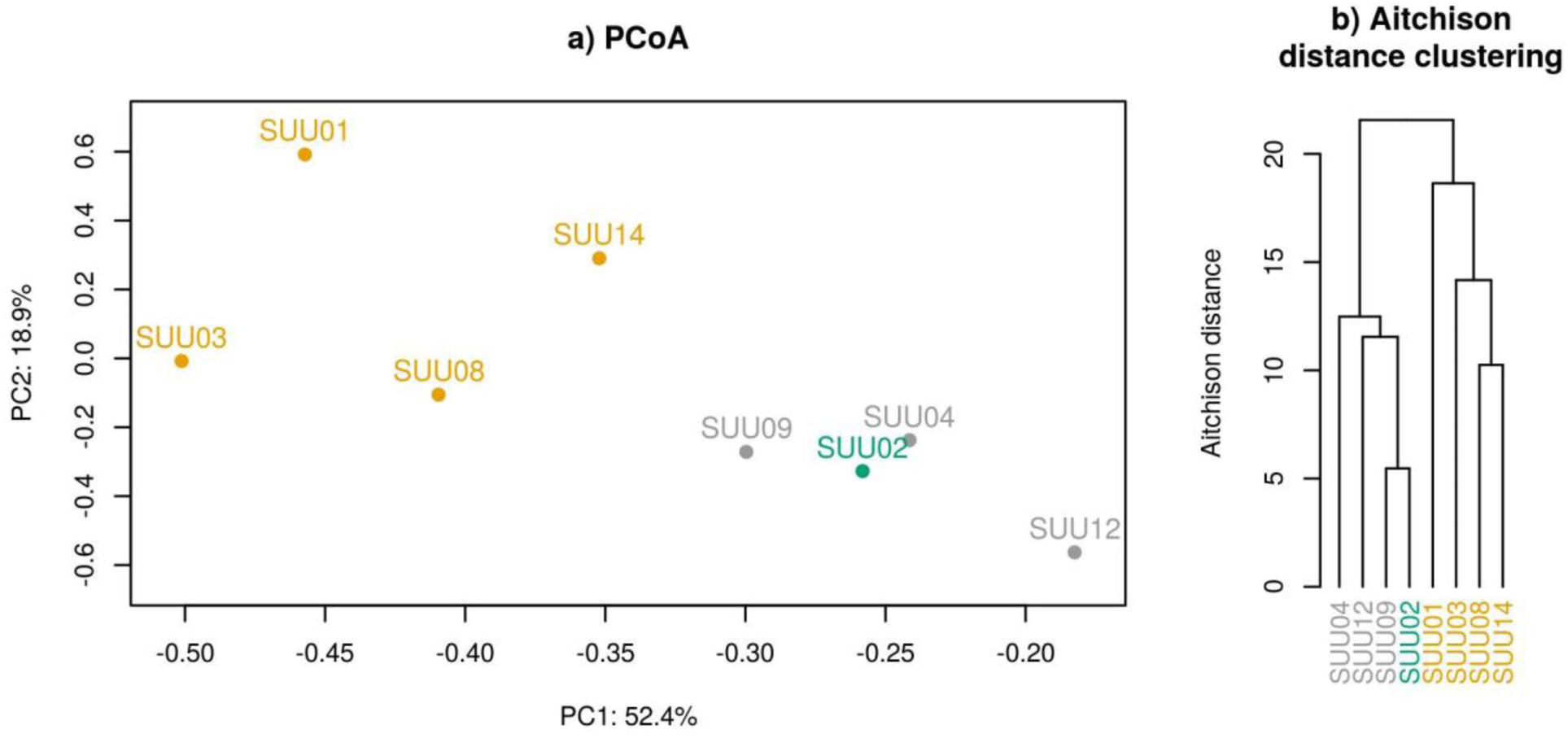
Visualization of the analysis of beta-diversity (Aitchison distance between samples): a) using the analysis of principal components in clr-coordinates, b) using clustering of samples according to the distance between them. The samples of the main group participants are marked in yellow, the samples of the comparison group are in gray. Sample SUU02, taken again from a patient from the main group, is highlighted in green.

In the second sample compared to the first one, the Shannon index increased from 3.84 to 4.4; the Chao1 index decreased from 137.34 to 99.76, respectively.

## Discussion

The majority of the microbial taxa detected in small intestine washing samples of the patients are observed in other studies about microbiota of different segments of the small intestine. Many of these are also known as common members of oral or colonic microbiota. Though being commensal, they may cause bacteremia and bacterial infection of organs if they get into the bloodstream. Several taxa were detected both in washing samples and in the negative control samples. Their presence in microbiota of patients with severe acute pancreatitis should be verified in further studies with larger sample size.

Comparison of patients with different disease severity shows decrease of relative abundance for taxa dwelling in the mucus layer in patients with higher scores according to integrative cumulative severity scales. Probably, this may be an evidence of association between acute pancreatitis complications and mucus layer thinning. The thinning may potentially lead to reduction of intestinal barrier function. If the reperfusion begins, it may also cause bacterial translocation through the damaged intestinal structures and development of systemic complications.

The second sample of a main group patient collected 24 hours after the end of SES enteral infusion manifested microbial composition that was more similar to the ones of the patients from the comparison group. The similarity was supported by beta diversity analysis, as well as by comparison of proportions of the major taxa that distinguished the main group from the comparison group. This may bear a therapeutic potential for future explorations of small intestinal microbiota composition correction for patients with acute severe pancreatitis.

The findings identified during this pilot study suggest that a study on extended cohort is required for validation. Particularly, a larger sample size is needed to make sound conclusions concerning the microbiome dynamics by comparing the paired samples before and after the SES. It is also important to investigate the composition of small intestinal microbiota in healthy subjects in order to assess the changes linked to the severe acute pancreatitis. Nevertheless, the pilot study results support the hypothesis of positive association between gut permeability and the disease severity and suggest that the further investigation of the problem may bring light to the understanding of pathogenic processes in the small intestine of patients with acute severe pancreatitis.

## Author contribution statement

VK, SP and PY contributed to the study concept and design; VK, AK, PY and SK contributed to the data acquisition; VK, VO and AT analyzed the data, contributed to the interpretation and wrote the manuscript.

## Additional information

The sample preparation, sequencing and data analysis of the microbiome samples were conducted by Knomics LLC as a part of a commercial contract for Vneshpromfarm LLC, manufacturer of the salt electrolyte solution (SES).

## Application

**FIGURE S1.**
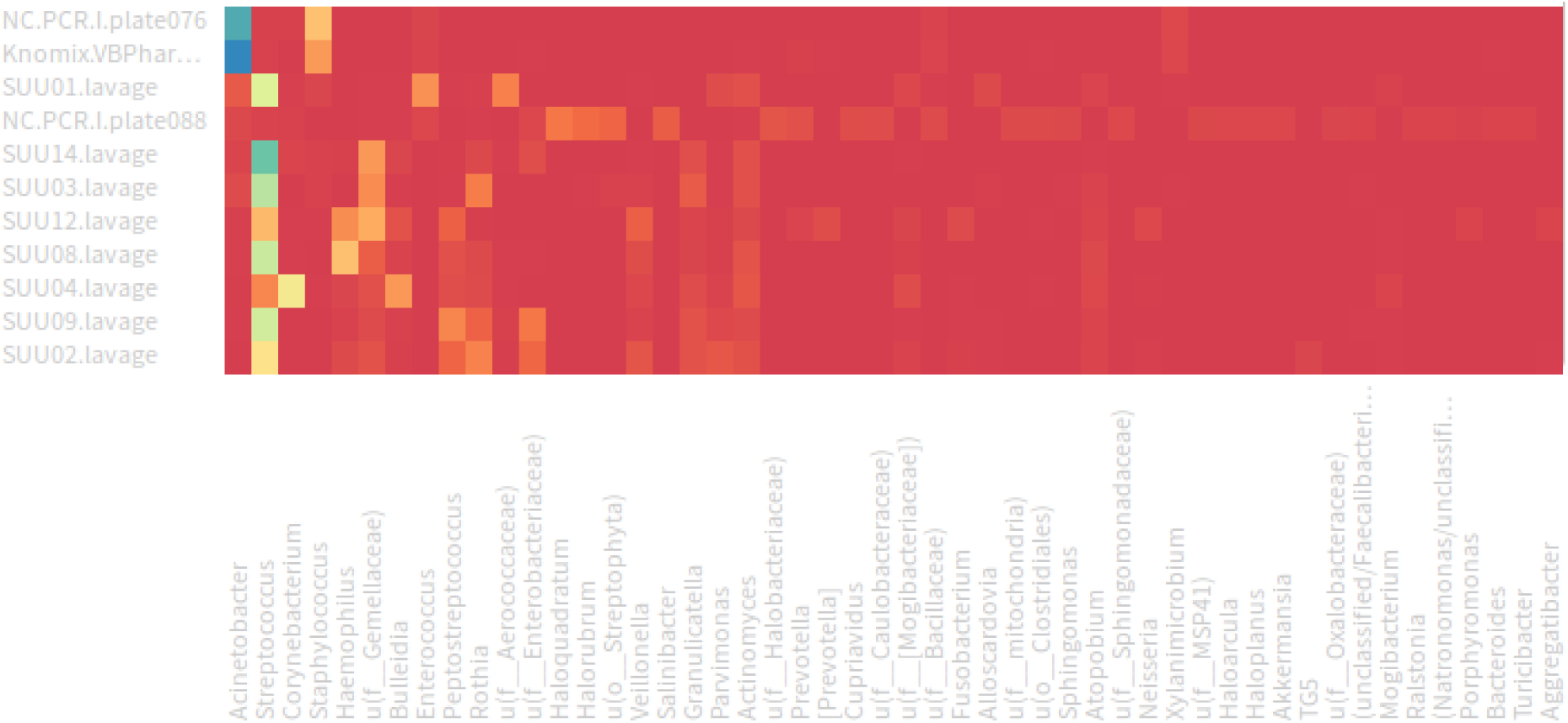
Heatmap of taxa representation (before manual removal of negative control reads).

